# Mortality gap continuum in severe mental illness: a nationwide study across healthcare and socioeconomic strata

**DOI:** 10.64898/2026.10.05.26364794

**Authors:** Gabriela Hjell, Lars Johan Hauge, Rune Johansen, Melanie Straiton, Per Henrik Zahl, Marit Haram, Maria Fernanda Vinueza Veloz, Øyvind Erik Næss, Carl Michael Baravelli, Ann Kristin Skrindo Knudsen, Kim Stene-Larsen, Anne Reneflot, Oleguer Plana-Ripoll, Benedicte Kirkøen, Martin Tesli

## Abstract

**Background:** A comprehensive overview of how premature mortality in severe mental illness varies by socioeconomic position and healthcare utilization is lacking.

**Methods:** We followed all Norwegian residents aged 30–79 years on January 1, 2008, through December 31, 2023. Nationwide registry linkage provided data on mental illness in primary and specialist care, educational attainment, and cause of death.

We estimated the mortality gap as excess life-years lost from age 30 for depression, bipolar disorder, and schizophrenia compared with the general population, stratified by sex, healthcare utilization, and educational attainment. All-cause, external-cause, and natural-cause mortality were assessed using competing risks.

**Results:** The cohort comprised 2,486,836 individuals, including 483,933 with depression, 40,125 with bipolar disorder, and 21,963 with schizophrenia. Among men, the mortality gap ranged from 1.50 year (95% CI 0.36–3.34) among those with depression, non-inpatient care utilization, and university education to 19.86 years (18.34–21.36) among those with schizophrenia, inpatient care utilization, and less than high school education. Among women, the corresponding range was no gap to 18.16 years (17.03–19.53). Mortality gaps were generally larger with lower educational attainment and inpatient care utilization and increased from depression to bipolar disorder to schizophrenia. This pattern was consistent across sexes, diagnoses, healthcare utilization levels, and educational attainment levels and was most pronounced for natural-cause mortality.

**Conclusions:** Premature mortality among people with severe mental illness forms a continuum across socioeconomic disadvantage, healthcare utilization, and diagnosis. Combining these measures can identify subgroups with the greatest mortality burden and may help target preventive interventions.

## Introduction

People with severe mental illness, including schizophrenia, bipolar disorder, and depressive disorder, have substantially increased risks of premature mortality (1–4). The premature mortality in severe mental illness arises from both natural causes comprising deaths due to diseases and medical conditions and from external causes comprising suicide, homicide, and deaths due to accidents (5–7). Meta-analyses show roughly doubled relative risks of all-cause mortality across schizophrenia (5), bipolar disorder (6), and depression (7), but relative risks are not suited to display how much lifespan is lost, or how much each cause of death contributes to that loss. The life-years lost (LYLs) method addresses both. It estimates the reduction in remaining life expectancy, and it can be decomposed by cause of death within a competing-risks framework (8, 9). Using this approach based on reduction in remaining life expectancy from the age at diagnosis, men with schizophrenia in the Danish population lost 12 years of lifespan, while women lost 9 years (1). The corresponding estimates were 8 years for men and 6 years for women with affective disorders. More recent data show the gap persisting or widening. In Scotland, the mortality gap for schizophrenia widened, rising to 12 lost years in men and 11 in women, with no improvement for bipolar disorder or depression (10). Updated Danish data for 2010–2023 show that standardized mortality ratios declined only modestly, with improvements confined to males and to some disorders such as depression, but not schizophrenia (11). Thus, despite ongoing initiatives to reduce the mortality gap in severe mental illness, this inequality remains a major public health concern (10–14). This underscores the need for more nuanced knowledge about mortality gap in different subgroups of severe mental illness, which can lead to improved targeted preventive strategies.

Severe mental disorders are heterogeneous and vary in clinical presentation and severity. In universal healthcare systems, individuals with more severe and recurrent symptoms often receive treatment in psychiatric inpatient units, whereas those with milder or more stable conditions may be treated in outpatient specialist mental health care and primary care (15). Although initial diagnostic assessments for schizophrenia and bipolar disorder typically occur in specialist mental health care (16, 17), a substantial proportion of individuals with these disorders are subsequently managed in primary care (18). In contrast, both diagnostic assessment and treatment of depressive disorder are typically organised within a stepped care framework, with increasing involvement of primary care, specialist outpatient services, and inpatient care as severity and healthcare needs escalate (19). Level of healthcare utilization is likely to matter for mortality, as estimates of premature mortality among people with severe mental illness were higher for those with inpatient care utilization than for those with outpatient care utilization (11, 20, 21). Still, no studies have estimated the mortality gap using the LYL method across different healthcare utilization levels within the same near universal healthcare system.

Disadvantaged socioeconomic position (SEP) is a determinant of adverse health outcomes, including increased risks of common mental disorders (22) and premature mortality in the general population (23, 24). Among people with mental disorders, SEP has been associated with elevated risks of all-cause mortality and natural-cause mortality (25). However, despite growing body of research examining the combination of mental disorders and different SEP levels linked to mortality (26, 27), understanding this association across different levels of healthcare utilization remains limited.

We therefore aimed to provide a comprehensive overview of how the premature mortality among people with severe mental disorders varies by level of healthcare utilization and socioeconomic position. Specifically, the main aim of this study was to determine the magnitude of mortality gaps among people with depression, bipolar disorder, and schizophrenia, separating those with inpatient care utilization versus those managed in outpatient or primary care, and further stratifying by educational attainment. To differentiate cases of depressive disorder that are typically more severe from milder presentations, we additionally aimed to assess a subgroup with depressive disorder with outpatient specialist care utilization and a subgroup with utilization of primary care only.

## Methods

### Study population and data sources

This population-based cohort study included all residents of Norway aged 30–79 years on Jan 1, 2008, who were followed up until Dec 31, 2023. The cohort selection flow chart is presented in **Supplementary figure 1**. The cohort is part of the SOC-MENT registry linkage, which contains sociodemographic and healthcare information retrieved from nationwide registries and linked at individual level using a unique personal identification number. Registry data sources encompassed the Norwegian Patient Registry (NPR) covering inpatient and outpatient contacts within specialist care from 2008 onwards, Norwegian Database for Control and Payment of Health Reimbursement (CPHR) that correspondingly covers contacts within primary care, Norwegian Cause of Death Registry, and Statistics Norway providing information on educational attainment. SOC-MENT has approval from The Regional Committees for Medical and Health Research Ethics (567529), and all study procedures were conducted in accordance with relevant guidelines, including the Declaration of Helsinki and the General Data Protection Regulation.

### Definitions of exposure strata

Information on specialist and primary care contacts for severe mental disorders diagnosed according to the International Classification of Diseases, 10th revision (ICD-10) or the International Classification of Primary Care, 2nd edition (ICPC-2) was obtained from the NPR and CPHR. Diagnoses of schizophrenia (ICD-10: F20; ICPC-2: P72), bipolar disorder (ICD-10: F30–31; ICPC-2: P73), and depression (ICD-10: F32–33; ICPC-2: P76) were retrieved, including information on the healthcare setting of all registry entries. For each disorder, we defined mutually exclusive levels of healthcare utilization: (1) inpatient care and (2) non-inpatient care including healthcare contacts across primary and specialist outpatient care without any inpatient contact. Any individual was considered unexposed until the first occurrence of the non-inpatient healthcare contact and, thereafter, considered as exposed to non-inpatient care until the first occurrence of the inpatient healthcare contact. In case that the first registry entry was an inpatient healthcare contact, the individual was considered unexposed until that timepoint and considered exposed to inpatient care thereafter.

SEP was operationalized using educational attainment at baseline (i.e., Jan 1, 2008), classified according to the International Standard Classification of Education (ISCED) as: lower than high school education (ISCED level 0-2), high school education (ISCED level 3-5), and university education (ISCED level 6-8) (28). To address potential age-related bias in SEP-stratified outcome estimates, we chose to study a cohort consisting of individuals aged 30 years or older as they represent a cohort with their own established SEP (26).

### Outcome definition

Information on all-cause mortality and the underlying cause of death was obtained from the Norwegian Cause of Death Registry. Causes of death were categorised as either (1) natural-cause mortality comprising deaths due to diseases and medical conditions (A00–R99) or (2) external-cause mortality comprising suicides (ICD-10: X60–X84, Y87·0), homicides (X85–Y09, Y87·1), and deaths due to accidents (V01–X59, Y85–Y86).

### Statistical analysis

Follow-up began on Jan 1, 2008, and ended at death, emigration, or Dec 31, 2023, whichever happened first. The mortality gap was estimated using the LYLs method (8, 9), based on calculations of remaining life expectancy from age 30 using age-specific mortality rates. LYLs quantified the average number of years lost between age 30 and 95, calculated as the difference between 95 and the estimated remaining life expectancy. The mortality gap was expressed as excess LYLs, comparing the LYLs estimate at age 30 for the category of interest to the LYLs estimate at age 30 in the general population of corresponding sex (i.e., the total cohort of men serving as a reference for calculations involving men in the specific disorder/healthcare-utilization/educational-attainment subgroup, and the total cohort of women serving as a reference for calculations involving women in the different subgroups). A competing risks model was applied to estimate excess LYLs attributable to natural and external causes of death (29). All estimates were reported with 95% CI derived using non-parametric bootstrapping with 1000 iterations. Data were analysed using the R software version 4.3.2, with excess LYLs analyses performed using the lillies package (9).

In the main analyses, we estimated mortality gaps for depression, bipolar disorder, and schizophrenia, stratified by level of healthcare utilization (inpatient, non-inpatient) and educational attainment (university, high school, lower than high school). Level of healthcare utilization was treated as a time-varying factor. The follow-up time of each individual was split into an unexposed part, non-inpatient part, and inpatient part according to the level of healthcare utilization, and each part was included in one of the mutually exclusive healthcare utilization categories (unexposed, non-inpatient, inpatient).

To differentiate cases of depressive disorder that are typically more severe from milder presentations, we estimated mortality gaps stratified by educational attainment separately in the subgroup with depressive disorder in outpatient specialist care (without any inpatient contact) and the subgroup with depressive disorder in primary care only.

Finally, to provide a broader context, we also performed analyses of 1) the mortality gap between the general population and each of the three disorders, 2) the mortality gap between the general population and each of the three disorders stratified by the level of healthcare utilization, and 3) the mortality gap between the general population and each of the three disorders stratified by educational attainment.

## Results

### Cohort description

The cohort consisted of 2,486,836 individuals (49.7% women), including 483,933 with depression (61.3% women), 40,125 with bipolar disorder (56.7% women), and 21,963 with schizophrenia (47.7% women). More detail is presented in **Table 1** and **Supplementary figure 2**.

**Table 1.** Median age at the start of the follow-up for depression, bipolar disorder, and schizophrenia by level of healthcare utilization and educational attainment.

|  |  | Non-inpatient care |  | Inpatient care |  |
| --- | --- | --- | --- | --- | --- |
|  |  | Men | Women | Men | Women |
| Depression | Lower than high school education | N= 46,269<br>50.4 years (17.6) | N=76,080<br>54.1 years (22.3) | N=8,012<br>54.5 years (22.8) | N=12,003<br>60.1 years (25.3) |
|  | High school education | N=83,104<br>51.5 years (17.6) | N=118,683<br>53.1 years (19.6) | N=11,335<br>57.2 years (21.3) | N=14,604<br>59.7 years (23.3) |
|  | University education | N=42,648<br>50.7 years (16.9) | N=83,940<br>48.0 years (15.5) | N=5,198<br>57.3 years (20.6) | N=7,348<br>53.3 years (19.4) |
| Bipolar disorder | Lower than high school education | N=3,568<br>48.0 years (15.5) | N=5,119<br>49.4 years (17.7) | N=1,755<br>51.6 years (16.8) | N=2,703<br>53.0 years (19.2) |
|  | High school education | N=6,127<br>49.6 years (16.3) | N=7,270<br>50.9 years (18.5) | N=2,989<br>54.3 years (18.2) | N=3,696<br>55.6 years (19.5) |
|  | University education | N=3,827<br>50.5 years (16.5) | N=5,491<br>47.5 years (15.2) | N=1,751<br>54.3 years (18.1) | N=2,344<br>50.7 years (17.6) |
| Schizophrenia | Lower than high school education | N=3,540<br>48.3 years (17.2) | N=3,289<br>55.2 years (21.9) | N=2,351<br>49.0 years (18.6) | N=1,768<br>54.7 years (20.1) |
|  | High school education | N=3,667<br>50.0 years (18.6) | N=3,359<br>56.5 years (18.7) | N=2,164<br>51.0 years (19.2) | N=1,623<br>56.8 years (19.5) |
|  | University education | N=1,420<br>49.0 years (17.9) | N=1,564<br>51.6 years (18.4) | N=730<br>50.1 years (18.3) | N=694<br>49.8 years (17.2) |

In parentheses: Interquartile range. Median age at the start of the follow-up in the general population was 50.8 years (21.4) among women and 50.0 years (20.9) among men.

### Main analyses

Among men, the mortality gap ranged from 1.50 excess LYL (95% CI 0.36 – 3.34) among those with depression, non-inpatient care utilization, and university educational attainment to 19.86 excess LYLs (18.34 – 21.36) among those with schizophrenia, inpatient care utilization, and less than high school educational attainment. Among women, the corresponding range was −0.63 (−0.87 – −0.34) excess LYLs to 18.16 excess LYLs (17.03 –19.53). Excess LYLs due to all-cause mortality was generally larger with lower educational attainment and inpatient care utilization and increased across diagnoses from depression to bipolar disorder to schizophrenia (**Figure 1** and **Supplementary table 1**). This overall pattern was consistent across sexes, diagnoses, healthcare utilization levels, and educational attainment levels.

**Figure 1.**
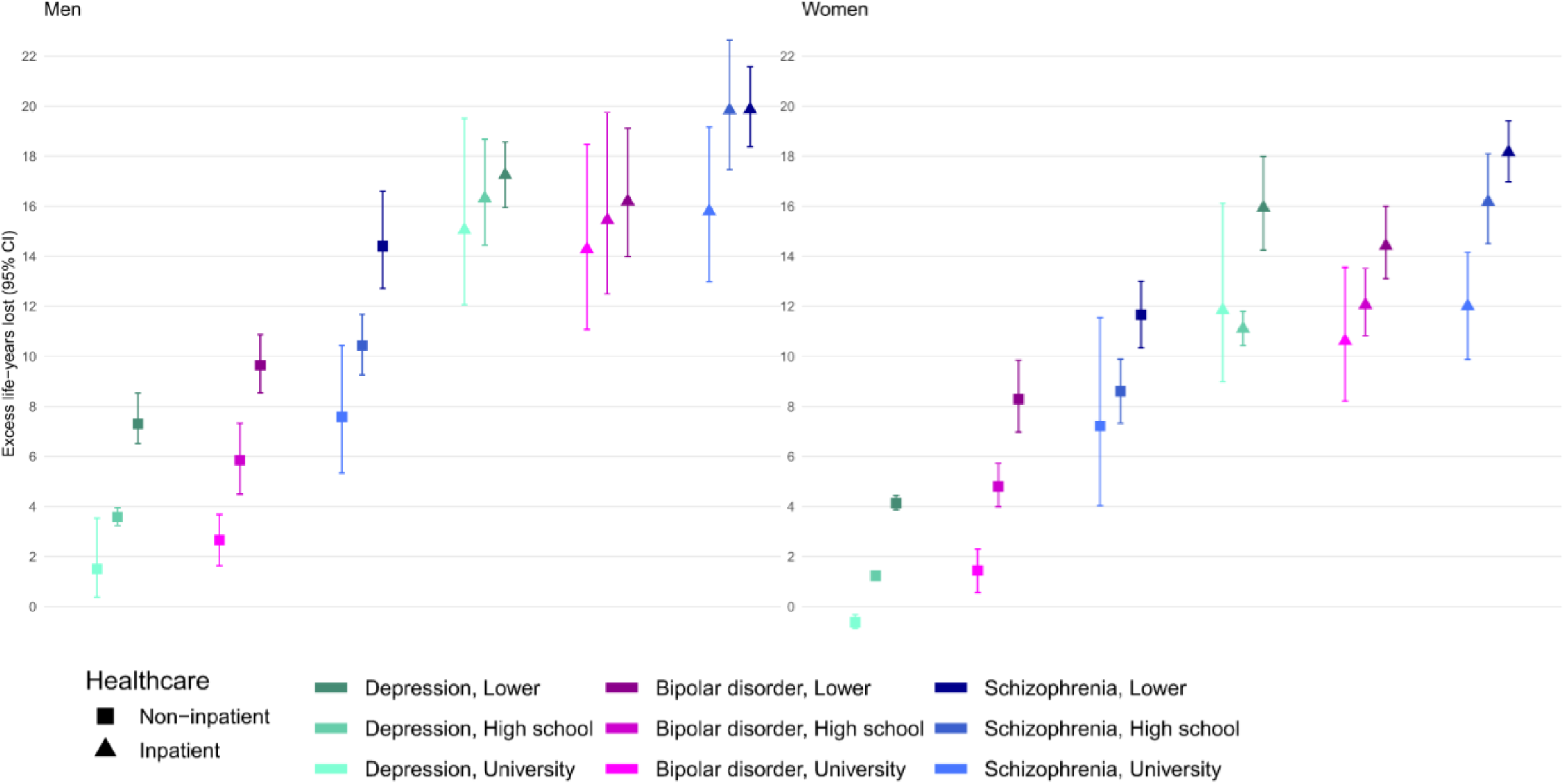
Excess life-years lost due to all-cause mortality among men and women with depression, bipolar disorder, and schizophrenia stratified by level of healthcare utilization and educational attainment. Excess life-years lost express the difference between remaining life expectancy at age 30 in the general population and remaining life expectancy at age 30 in the displayed stratified patient groups.

Across both sexes, excess LYLs due to natural-cause mortality was also generally larger with lower educational attainment and inpatient care utilization and increased across diagnoses from depression to bipolar disorder to schizophrenia. The range of excess LYLs due to natural causes was up to 12.04 (10.53 – 13.75) among men and up to 14.39 (13.11 –15.73) among women. For excess LYLs due to external causes, the range was up to 12.67 (8.12 – 17.94) among men and up to 8.69 (5.00 – 14.06) among women. In contrast, there were no clear patterns in mortality gap due to external causes according to disorder or educational attainment, and larger gaps due to external-cause mortality among people with inpatient care utilization compared to inpatient care utilization were only seen in depression and bipolar disorder (**Figure 2, Supplementary table 1**).

**Figure 2.**
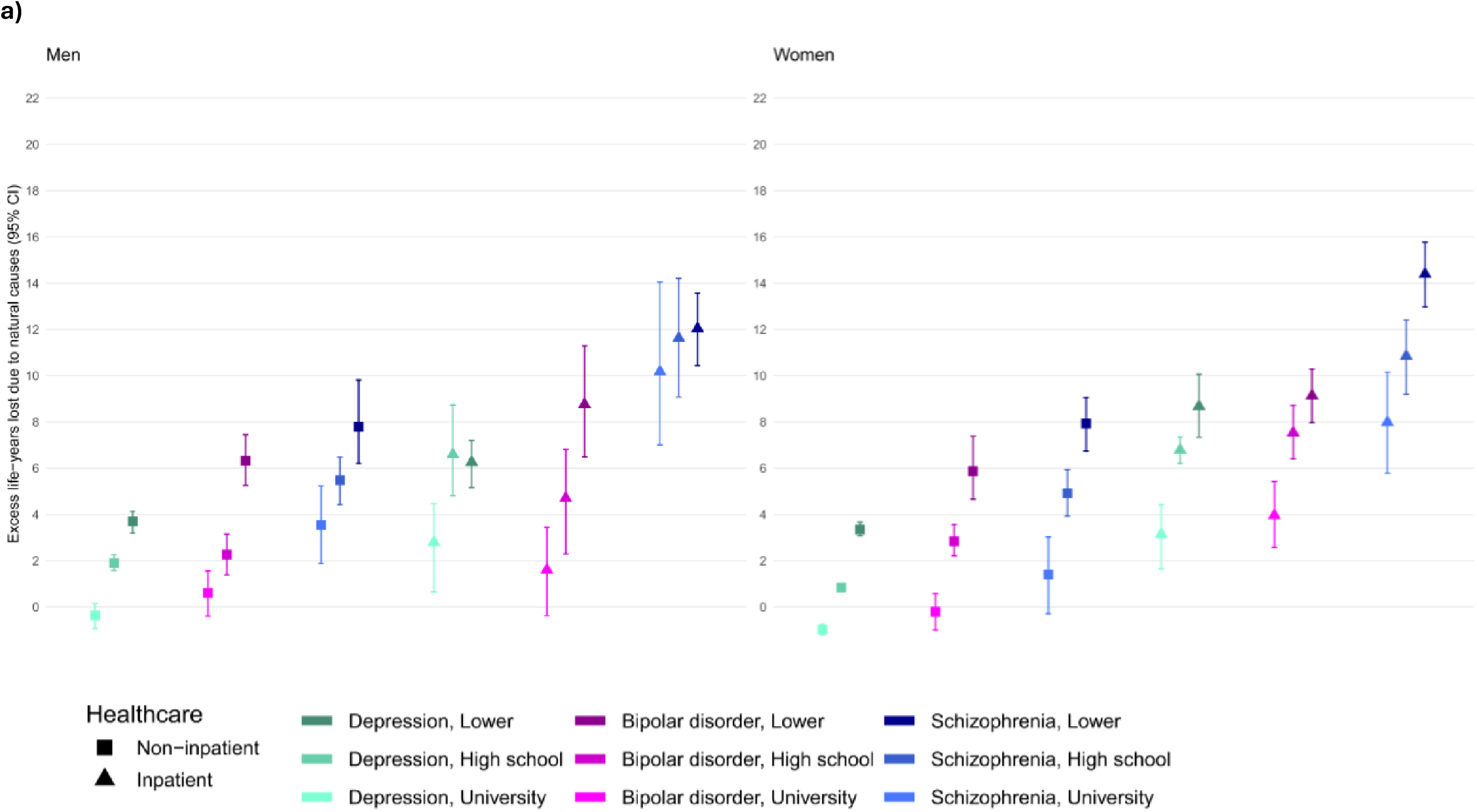
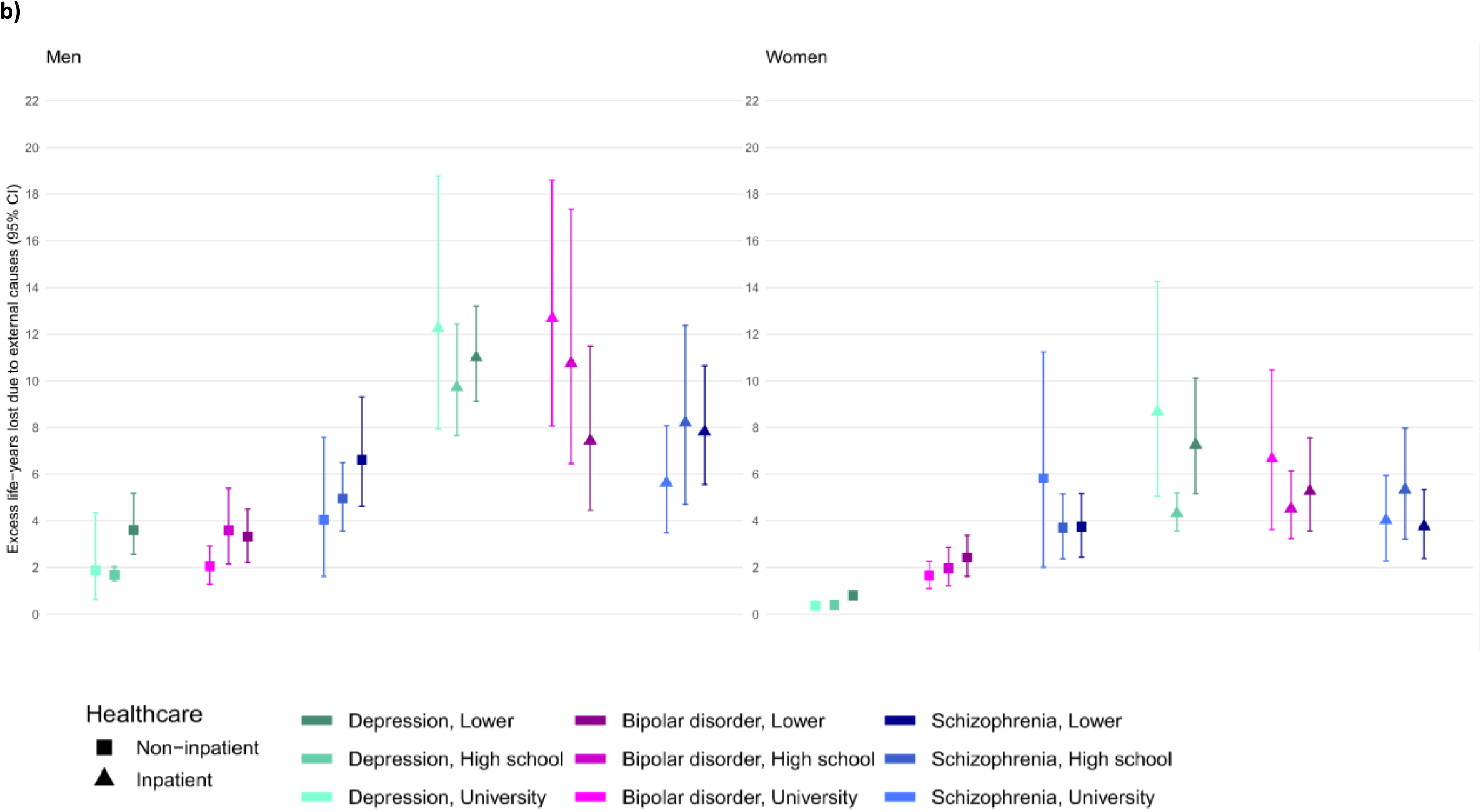
Excess life-years lost due to a) natural-cause and b) external-cause mortality among women and men with depression, bipolar disorder, and schizophrenia stratified by level of healthcare utilization and educational attainment. Excess life-years lost express the reduction in life expectancy at age 30 due to a) natural-cause mortality and b) external-cause mortality among the displayed stratified patient groups compared to the general population.

### Secondary analysis of excess LYLs in depressive disorder

Across all educational attainment levels, men with depression in outpatient specialist care had similar excess LYLs due to all-cause mortality as those in primary care only. For example, the gap due to all-cause mortality among men with high school educational attainment and depression with outpatient specialist care utilization was 4.48 excess LYLs (3.99 – 5.00), while the gap among those with primary care utilization only was 3.36 excess LYL (2.96 – 3.85). Corresponding estimates for women were 2.69 excess LYLs (2.29 – 3.14) and 0.94 excess LYL (0.76 – 1.12). The pattern of larger excess LYLs among women in outpatient specialist care than those in primary care only was seen across all educational attainment levels (**Supplementary table 2**). The gap due to all-cause mortality among men with depression in outpatient specialist care and university educational attainment was 1.69 excess LYLs (0.86 – 2.70) and the corresponding gap among women was 0.58 excess LYL (0.00 – 1.20).

### Additional analyses

Without further stratification by educational attainment or healthcare utilization, excess LYLs were smallest for depression, intermediate for bipolar disorder, and largest for schizophrenia (**Supplementary table 3**). Compared with men and women in the general population, the gap due to all-cause mortality was 5.89 excess LYLs (95% CI 5.51 – 6.41) among men with depression and 3.11 excess LYLs (2.97 – 3.27) among women with depression. For bipolar disorder, the corresponding estimates were 9.29 (8.63 – 9.99) and 8.09 (7.61 – 8.59) excess LYLs, and for schizophrenia 15.45 (14.59 – 16.38) and 12.73 (12.04 – 13.54) excess LYLs.

When stratified by the level of healthcare utilization without further stratification by educational attainment, there was a larger gap due to all-cause and natural-cause mortality among people with each of the three disorders with inpatient care utilization than among those with non-inpatient care utilization (**Supplementary table 4**). The corresponding gap due to external-cause mortality was larger among people with depression with inpatient care utilization and among people with bipolar disorder with inpatient care utilization than among those with non-inpatient care utilization. The corresponding estimates were similar between the healthcare utilization levels in schizophrenia.

When stratified by educational attainment without further stratification by the level of healthcare utilization, the gap due to all-cause and natural-cause mortality was generally larger with lower educational attainment in each of the three disorders (**Supplementary table 2**). The corresponding gaps due to external-cause mortality in each of the three disorders were similar between the educational attainment levels.

## Discussion

In this population-based study providing a comprehensive overview of premature mortality in severe mental illness by sex, diagnosis, socioeconomic position and healthcare utilization, we identified a mortality gap ranging from no gap to 20 excess LYLs. The mortality gap was generally larger with lower educational attainment and among people with inpatient care utilization and increased from depression to bipolar disorder to schizophrenia. These gradients were pronounced for all-cause and natural-cause mortality and consistent across sexes and within diagnostic, healthcare utilization, and educational attainment strata. Excess LYLs due to external-cause mortality was also substantial but showed no clear pattern.

Results from our main analyses of mortality gap among people with severe mental illness from age 30 showed none to 20 excess LYLs across diagnostic, healthcare utilization, and educational attainment strata. While evidence on magnitude of premature mortality in severe mental illness vary widely depending on statistical methods applied and populations studied, the estimates fall within the range of excess LYLs identified in our study (2, 3). Results of our analyses of mortality gap from age 30 without stratification by level of healthcare utilization and educational attainment that were performed to provide a broader context showed 15 excess LYLs among men and 13 excess LYLs among women with schizophrenia, 9 excess LYLs among men and 8 excess LYLs among women with bipolar disorder, and 6 excess LYLs among men and 3 excess LYLs among women with depression. This is broadly consistent with findings from population-based studies that used similar methodology (1, 10, 30–35) and in line with meta-analytic evidence of substantially reduced life expectancy among individuals with severe mental disorders (2–4). Our results showed mortality gaps of distinct magnitudes between the three disorders. When stratified by level of healthcare utilization and educational attainment, these differences between disorders transformed into a continuum, indicating a substantial heterogeneity within the groups.

In line with the literature showing excess mortality in severe mental disorders due to natural causes such as cardiovascular disease, neoplasms, respiratory disease, gastrointestinal disease, and metabolic diseases (1, 36, 37), we found substantial excess LYLs due to natural causes of death. Further, in line with evidence of premature mortality due to external causes such as accidents and suicide (1, 2, 36), we also found excess LYLs due to external causes of death, supporting that external causes of death contribute to mortality gap also after age 30.

While confirming the well-established evidence of premature mortality in severe mental illness derived from specialist care (2, 3), our findings also support a broader generalisability of this knowledge by demonstrating a mortality gap even when primary care is included. Initial diagnostic assessments for schizophrenia and bipolar disorder typically occur in specialist mental health care (16, 17). However, a substantial proportion of individuals with these disorders are subsequently managed in primary care (18). In previous population-based nationwide studies of mortality gap in severe mental illness, the time window between the earliest possibility to capture an outpatient care contact and the end of the follow-up ranged from 10 (30, 31) to 20 (1) years, which is shorter than the typical timeframe between the age of illness onset and the age at death (38). Thus, our findings based on assessments comprising primary care settings also account for mortality among individuals who do not require specialist mental health care during later phases of their illness course.

We found a gradient in all-cause and natural-cause mortality showing excess LYLs that was generally larger with lower educational attainment and among people with inpatient care utilization and increased from depression to bipolar disorder to schizophrenia. In contrast, excess LYLs due to external-cause mortality showed no clear pattern. By quantifying the effects of diagnostic, healthcare utilization, and educational attainment strata in an integrated and detailed manner, these findings extend the evidence on how clinical and SEP-related characteristics among people with severe mental illness jointly contribute to the mortality gap (26, 39). For example, among men with non-inpatient care utilization, all-cause mortality gap estimates for both those with schizophrenia and high school educational attainment and those with bipolar disorder and lower than high school educational attainment were 10 excess LYLs. This demonstrates how the mortality gap continuum encompasses clinical and SEP-related differences in premature mortality, while some of the granular estimates are of similar magnitudes across the diagnoses.

We found an educational gradient in mortality gap that was largely driven by excess LYLs due to natural causes. This aligns with a larger socioeconomic disparity in mortality rates due to natural but not external causes among people with severe mental illness (26) and corresponding larger socioeconomic disparities in mortality due to several specific natural causes of death, including cardiometabolic and respiratory diseases (27). Our results of educational gradient in the mortality gap in a population with severe mental illness may also reflect reverse causality and potentially illness severity, as individuals suffering from severe mental illness may be prevented from fulfilling their educational potential (40).

We found a larger gap due to all-cause, natural-cause, and external-cause mortality among people with bipolar disorder and depression with inpatient care utilization. In contrast, while people with schizophrenia with inpatient care utilization exhibited larger gap due to all-cause and natural-cause mortality compared to those with non-inpatient care utilization, we found no differences in healthcare utilization level related gap in mortality due to external causes of death. This is largely consistent with indications of higher mortality risk (20) and higher standardized mortality ratios (11, 21) among people with severe mental illness and history of hospitalisation. These findings demonstrate that the clinical heterogeneity in severe mental disorders is linked to corresponding heterogeneity in premature mortality. This further indicates that history of hospitalisation may serve as a risk marker of premature mortality due to natural causes of death across the three disorders and premature mortality due to external causes of death among people with depression and bipolar disorder.

Women with depression in outpatient specialist care had somewhat larger mortality gap compared to those in primary care only, while there were no differences in mortality gaps among men. This is in line with substantially larger difference between mortality rates among women with depression in outpatient and primary care compared to corresponding difference among men (39). One possible explanation might be that higher proportion of women than men with depression in primary care suffer from a clinical condition associated with milder adverse outcome characteristics, potentially associated with a lower threshold for seeking health care for depressive symptoms among women compared to men (41).

This study has several strengths. First, the study include data from a large nation-wide registry linkage covering both specialist and primary care. Second, this study contributes, for the first time, with population-based estimates of the mortality gap in severe mental illness from Norway. Third, the study provides a comprehensive investigation of how the mortality gap varies according to level of healthcare utilization and SEP. Fourth, differentiating external and natural mortality provides substantially more insights compared to estimates due to all-cause mortality alone. This study also has some limitations. First, estimating excess LYLs from 30 years may have led to somewhat underestimated premature mortality since the estimates are by design conditioned on survival until age 30. Second, despite use of data from nationwide registries, risk of misclassification of healthcare utilization category cannot be fully ruled out since the study design did not capture potential healthcare contacts that occurred before start of the follow-up. Third, our findings may not directly generalise to countries without near-universal access to medical care and education, particularly considering use of educational attainment and healthcare utilization level as distinct subgroup characteristics. Furthermore, our findings may not generalise to population of people with severe mental illness that never engage with the healthcare system.

To conclude, our findings indicate that premature mortality among people with severe mental illness forms a continuum across socioeconomic, healthcare utilization, and diagnostic strata and ranges from no gap to 20 years. The findings further indicate that lower educational attainment, inpatient care utilization, and diagnostic category jointly contribute to the pronounced mortality gap due to natural causes, whereas the premature mortality due to external causes, which is also substantial, exhibits less clear pattern. The present granular overview suggests that combining these clinical, healthcare, and sociodemographic metrics can identify subgroups with the greatest mortality burden and may help target preventive interventions.

## Data Availability

Researchers can apply for access to health registry data including CPHR (https://www.fhi.no/he/kpr/) and NPR (https://www.fhi.no/he/npr/sok-om-data-fra-npr/).

