## Supplementary information for "Mortality gap continuum in severe mental illness: a nationwide study across healthcare and socioeconomic strata"

**Supplementary figure 1.** The cohort selection flow chart.

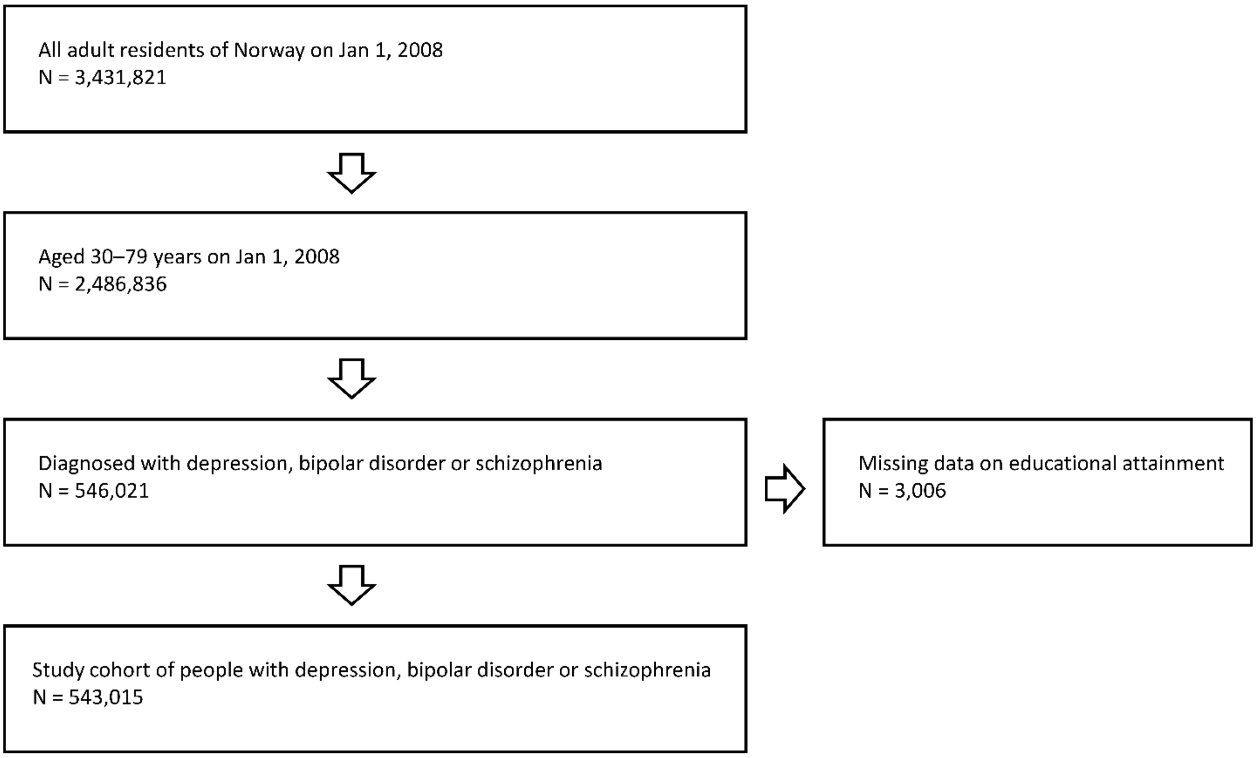

**Supplementary figure 2.** Age at the end of the follow by sex, diagnosis, level of healthcare utilization, and educational attainment.

W = Women, M = Men, scz = Schizophrenia, bd = Bipolar disorder, mdd = Depression, Inpatient = Inpatient care utilization, NonInpatient = Healthcare utilization across primary and specialist outpatient care without any inpatient contact, Uni = University education, High = High school education, Low = Lower than high school education.

N=694

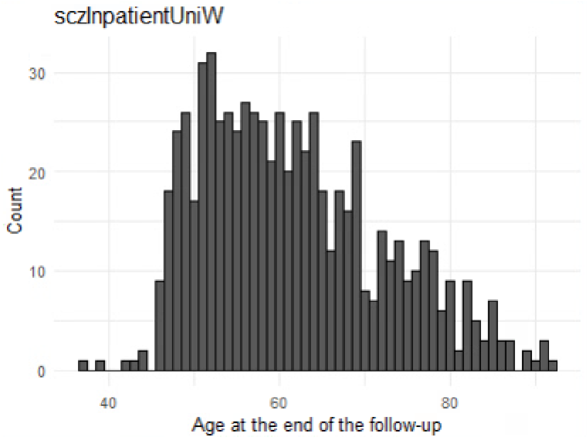

N=730

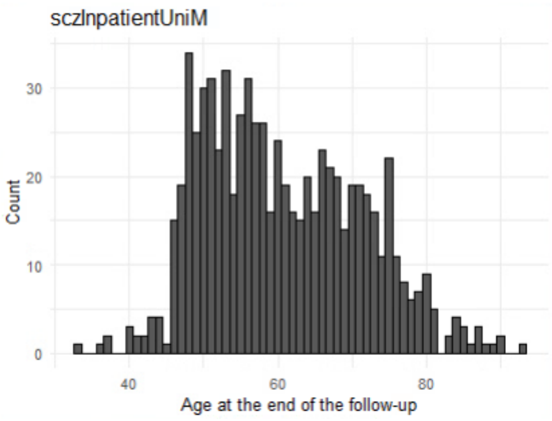

N=1623

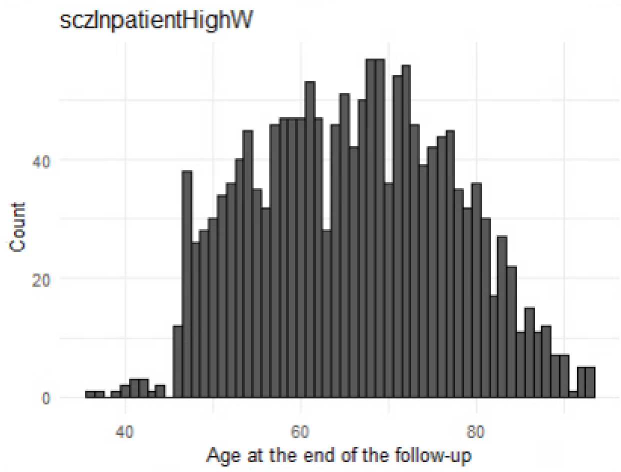

N=2164

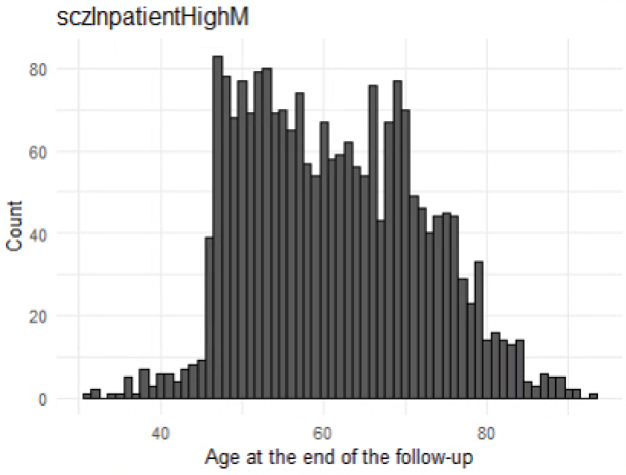

N=1768

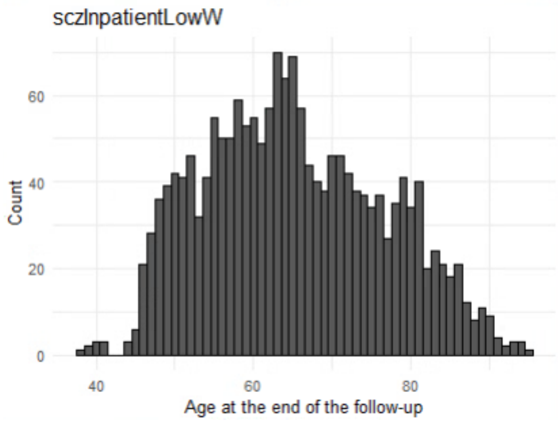

N=2351

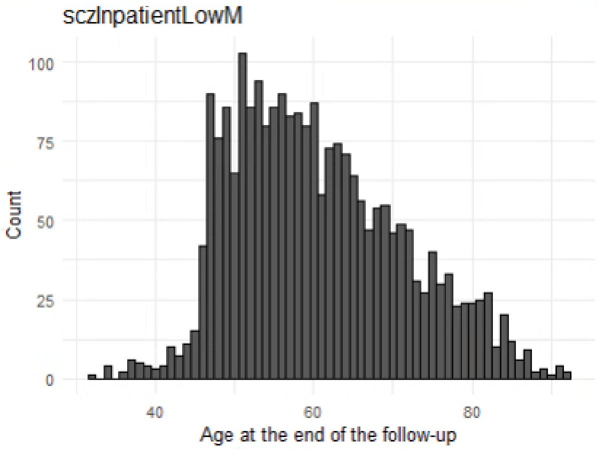

N=1564

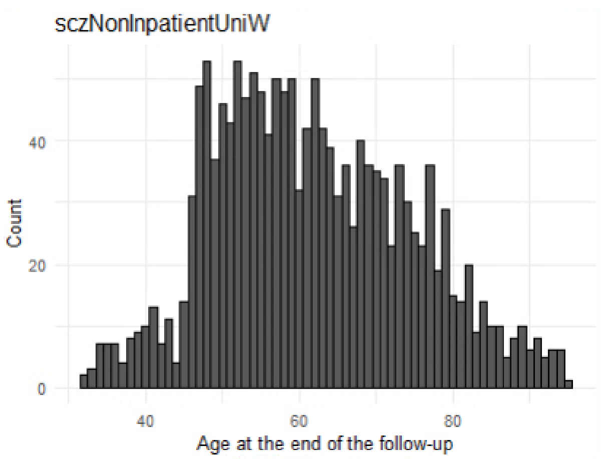

N=1420

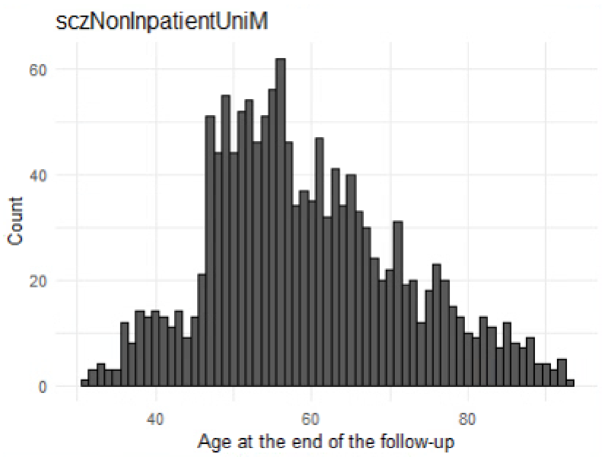

N=3359

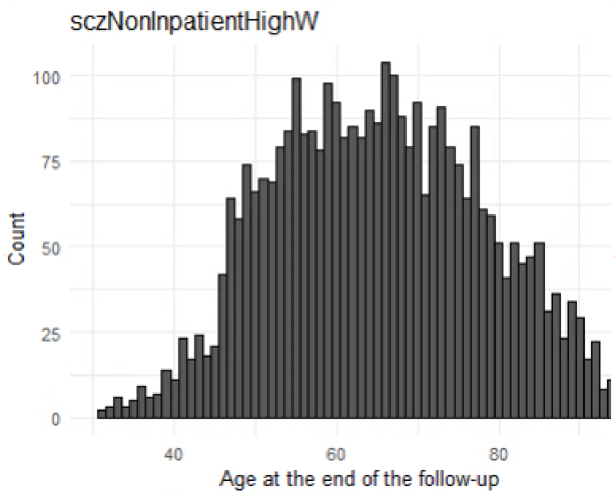

N=3667

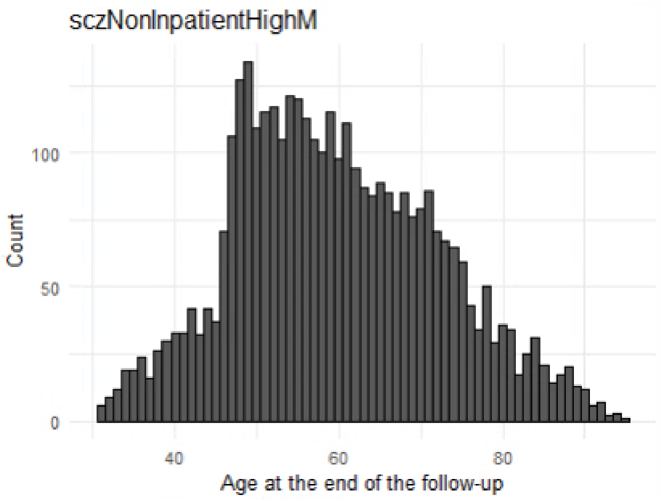

N=3289

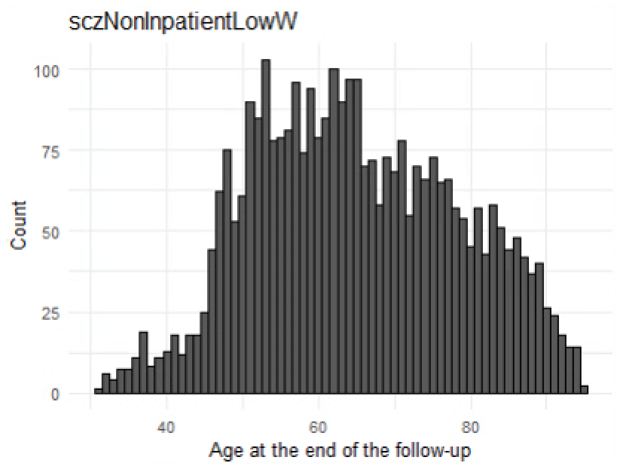

N=3540

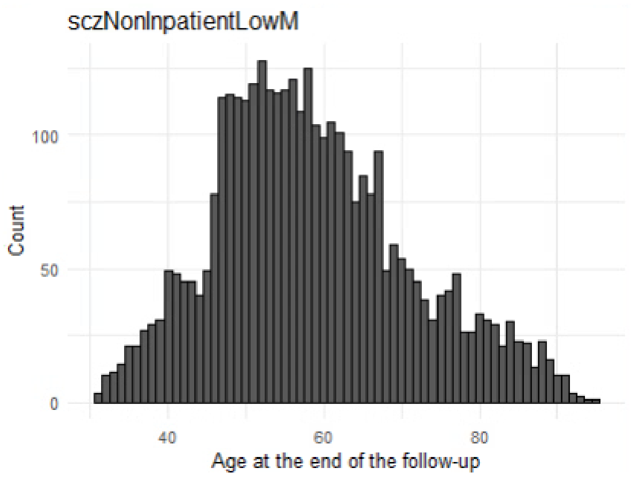

N=2344

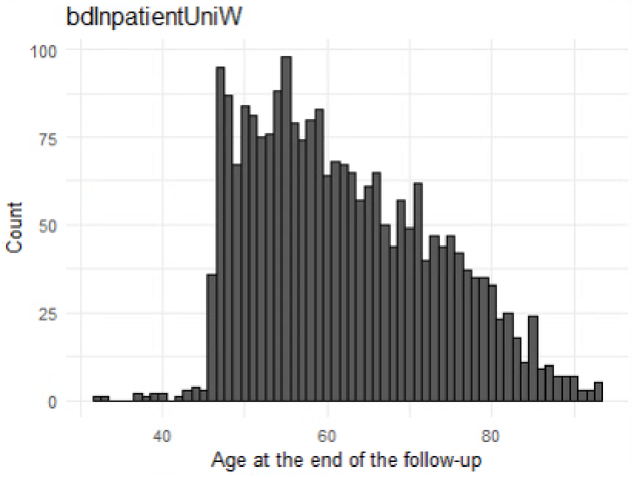

N=1751

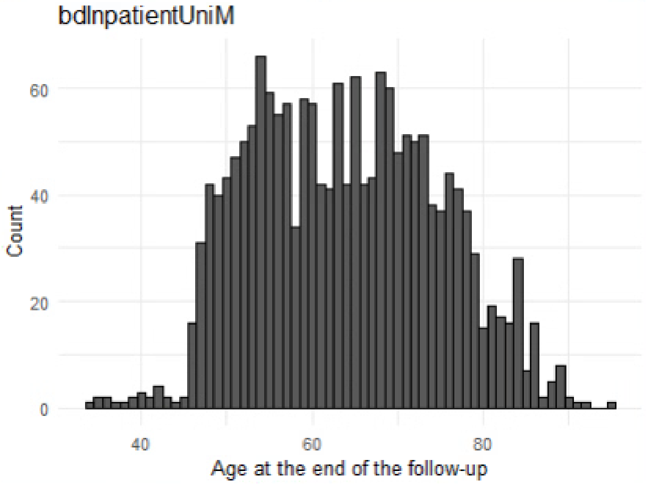

N=3696

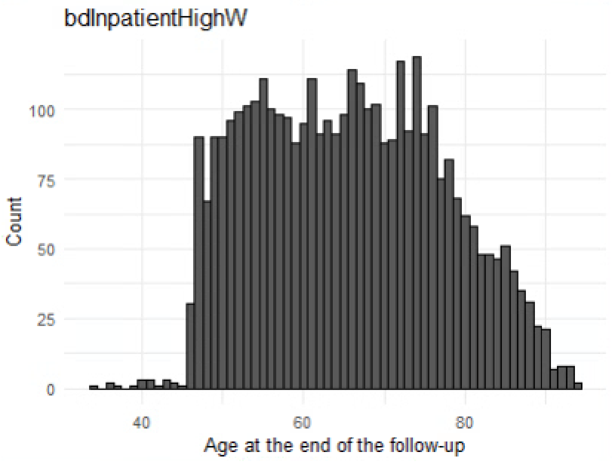

N=2989

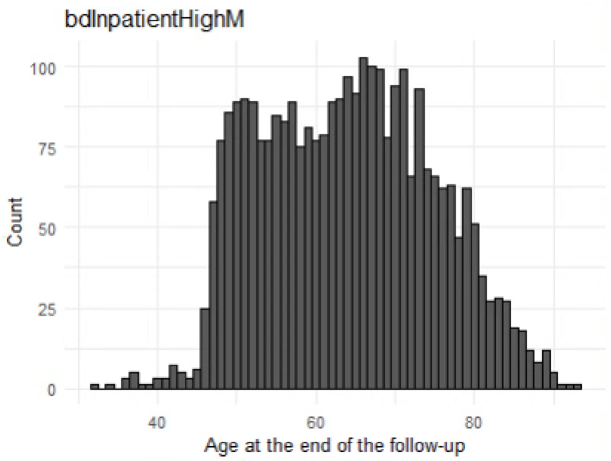

N=2703

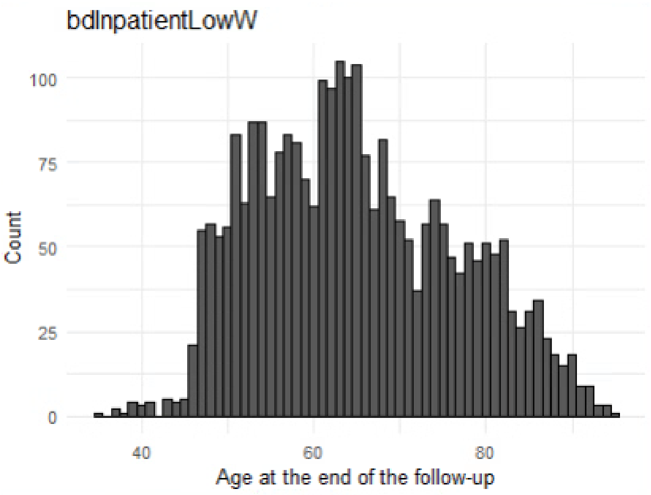

N=1755

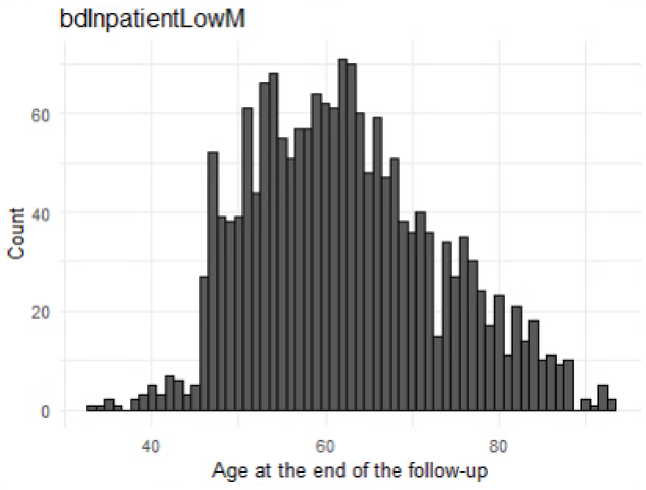

N=5491

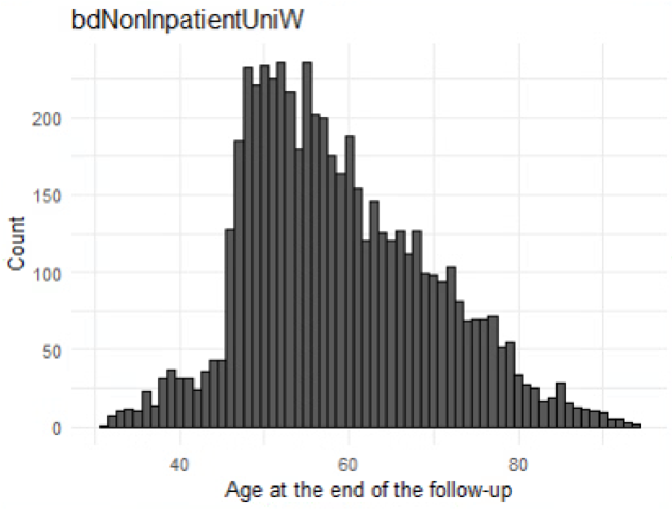

N=3827

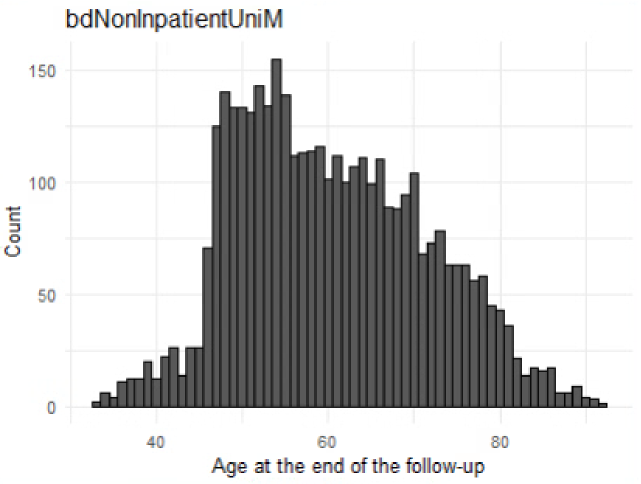

N=7270

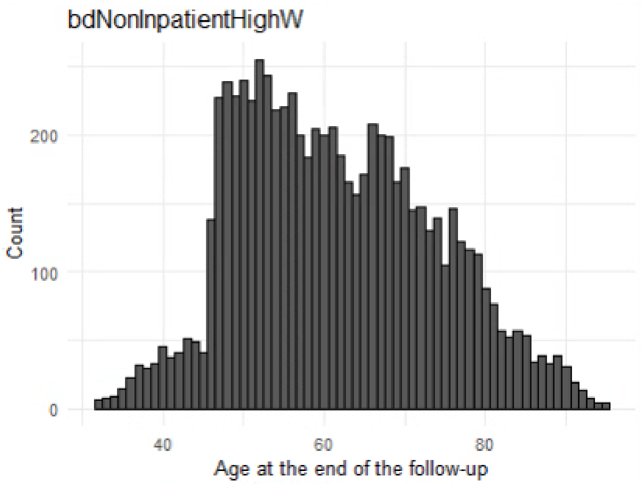

N=6127

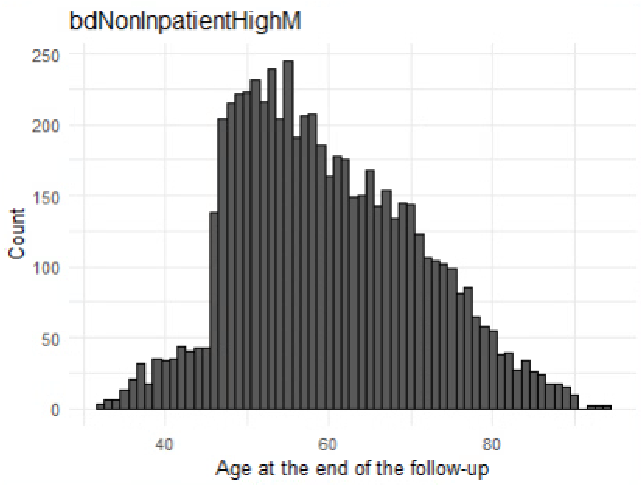

N=5119

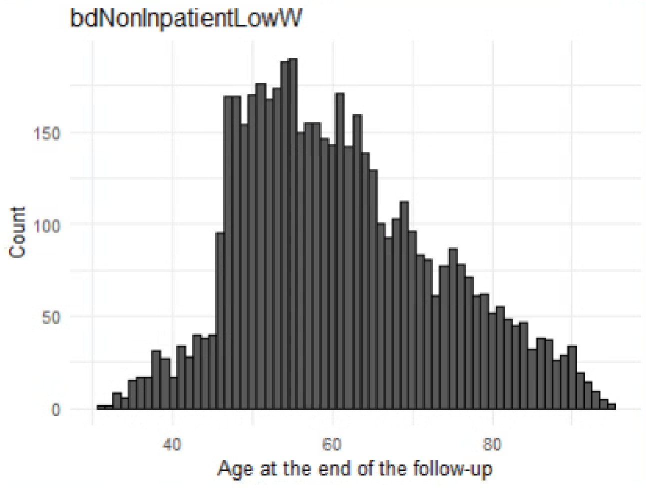

N=3568

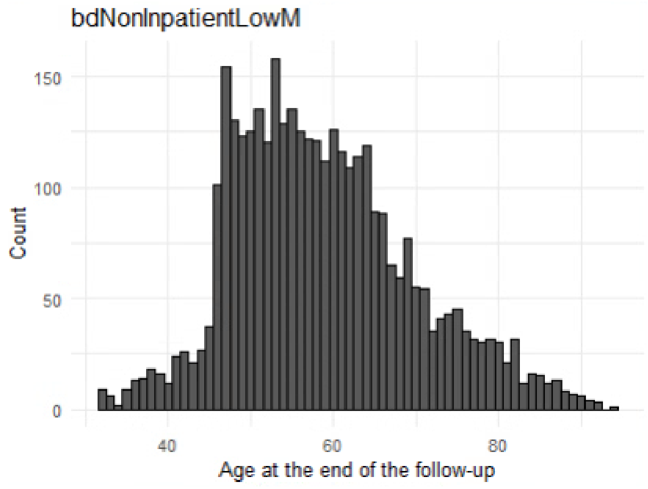

N=7348

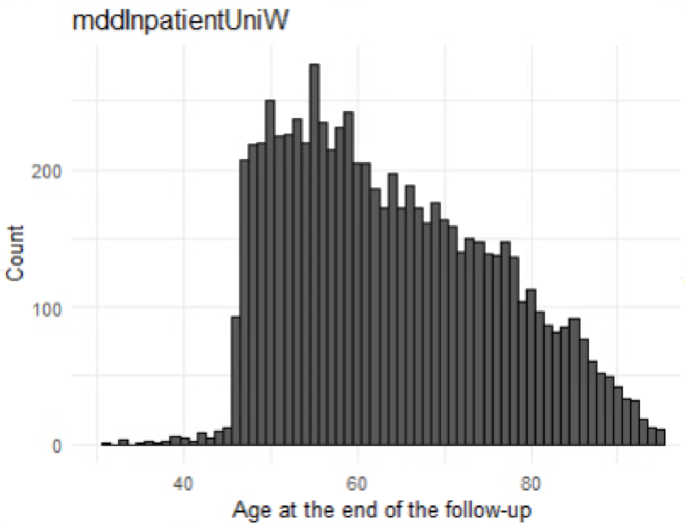

N=5198

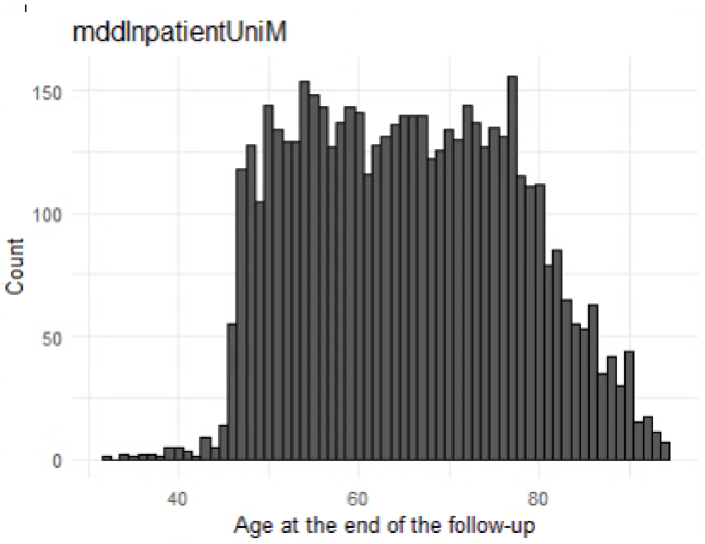

N=14604

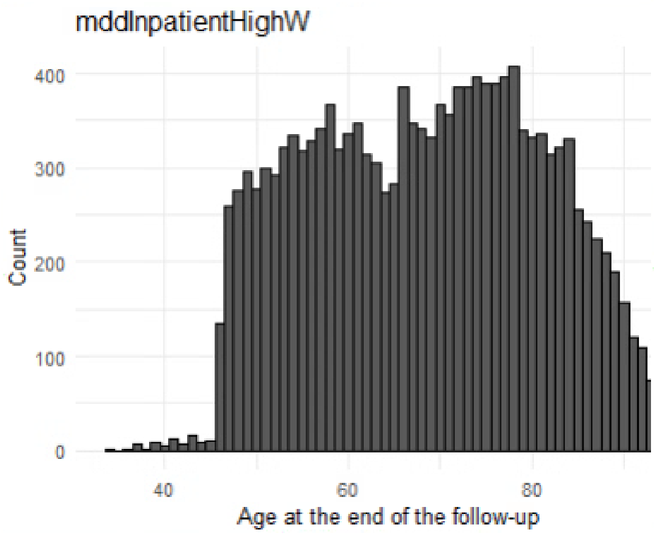

N=11335

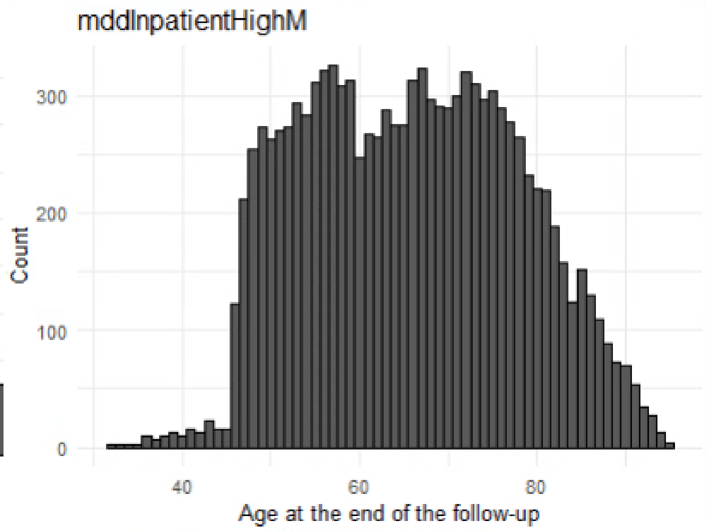

N=12003

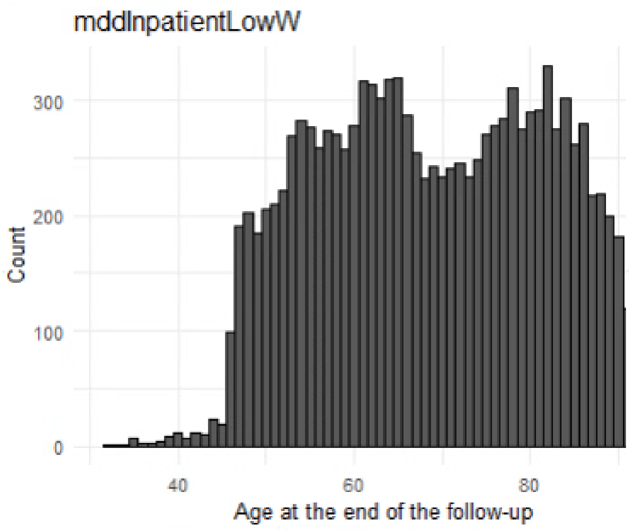

N=8012

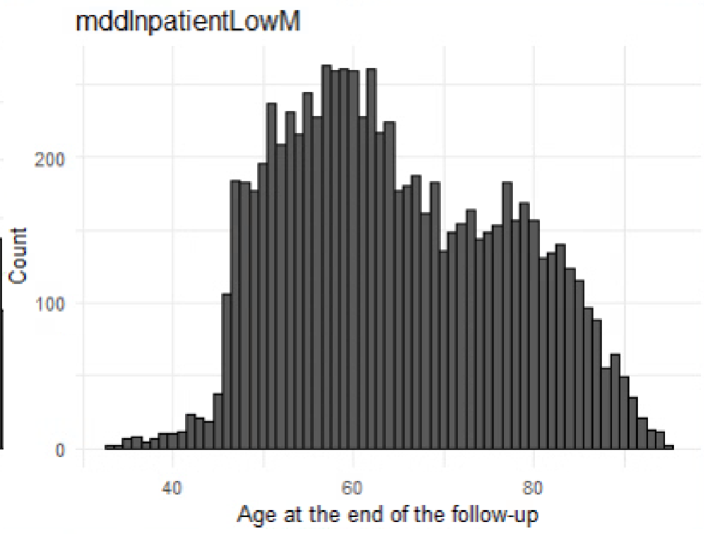

N=83940

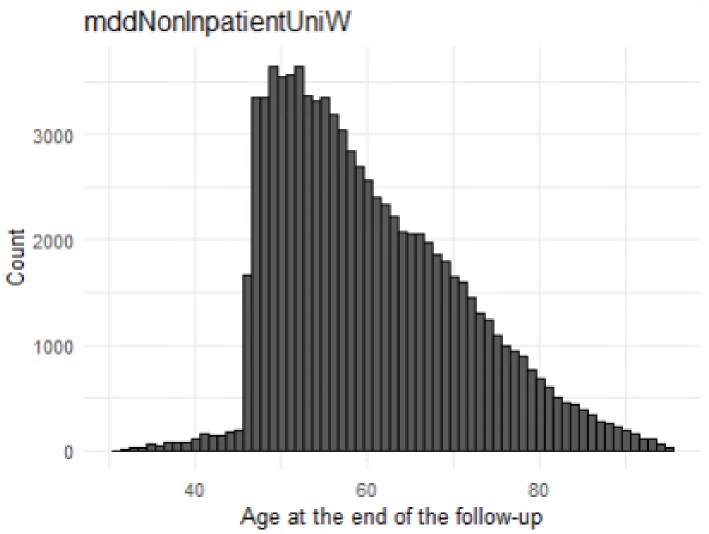

N=42648

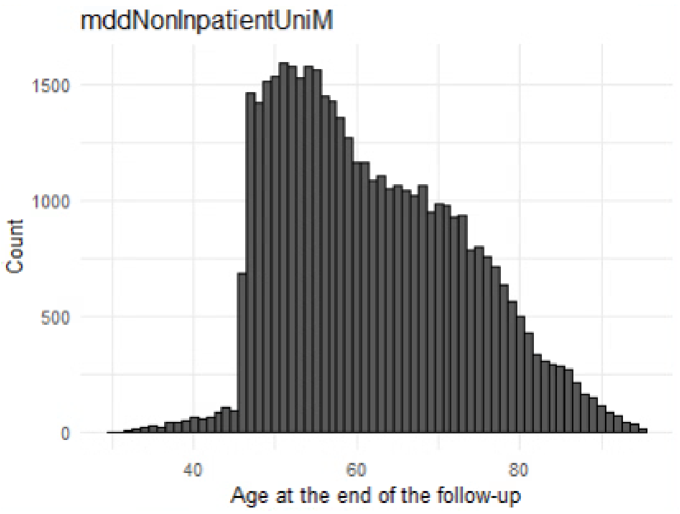

N=118683

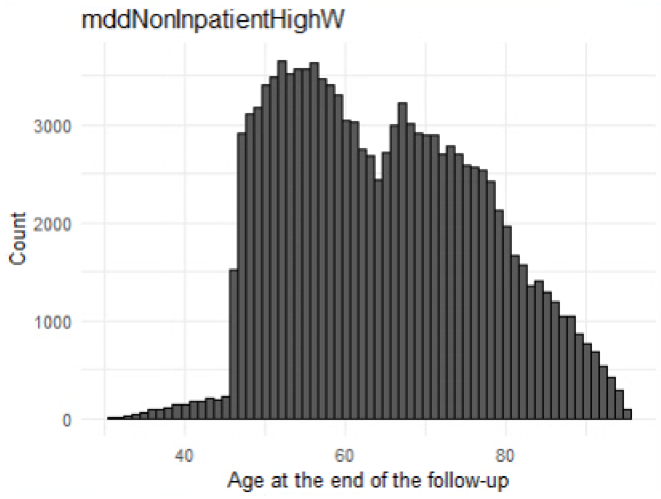

N=83104

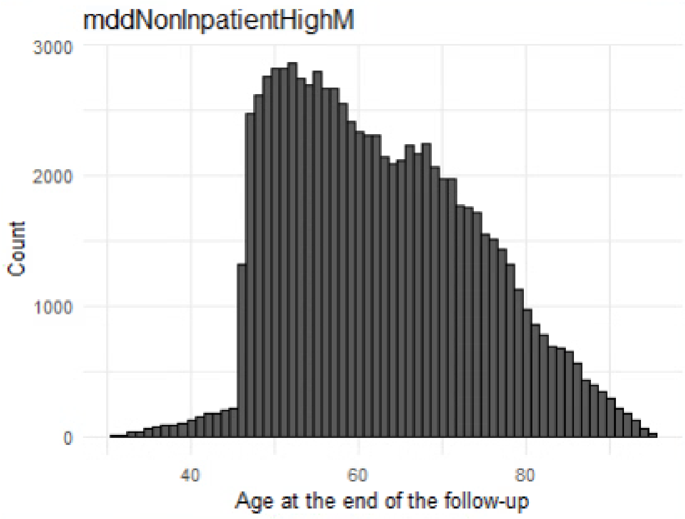

N=76080

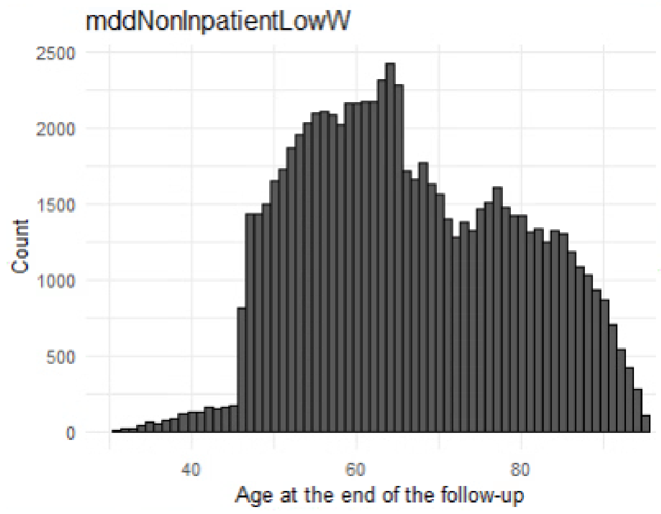

N=46269

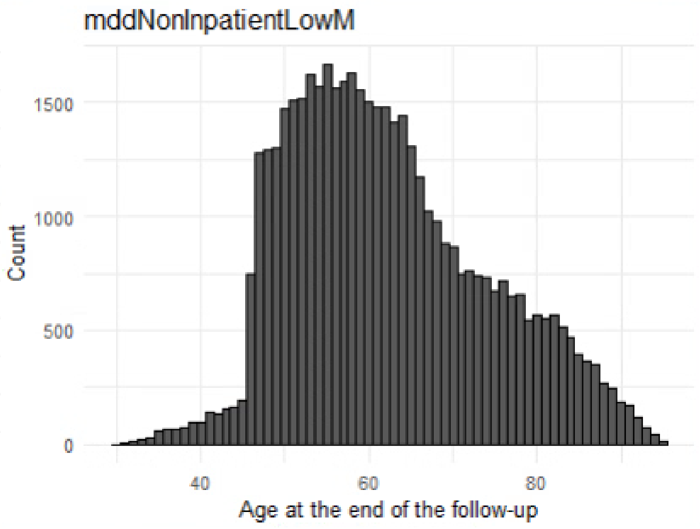

**Supplementary table 1.** Excess life-years lost from age 30 in a) women and b) men with schizophrenia, bipolar disorder, and depression according to level of healthcare utilization and educational attainment.

a) Women

| | | | |
|---|---|---|---|
| Schizophrenia | | | |
| | | Inpatient | Non-inpatient |
| - Lower than high school | Total cause of death<br>Natural<br>External | 18.16 (17.03 – 19.53)<br>14.39 (13.11 – 15.73)<br>3.77 (2.46 – 5.35) | 11.67 (10.43 – 13.09)<br>7.92 (6.76 – 9.03)<br>3.75 (2.50 – 5.46) |
| - High school | Total cause of death<br>Natural<br>External | 16.18 (14.44 – 18.16)<br>10.85 (9.31 – 12.46)<br>5.33 (3.15 – 7.99) | 8.61 (7.37 – 9.85)<br>4.91 (3.90 – 5.85)<br>3.70 (2.37 – 5.30) |
| - University | Total cause of death<br>Natural<br>External<br>Life expectancy | 12.01 (9.91 – 14.24)<br>7.98 (5.77 – 10.18)<br>4.02 (2.31 – 5.88) | 7.22 (4.03 – 12.24)<br>1.41 (-0.29 – 2.96)<br>5.81 (2.06 – 11.93) |
| Bipolar disorder | | | |
| | | Inpatient | Non-inpatient |
| - Lower than high school | Total cause of death<br>Natural<br>External | 14.42 (13.07 – 16.00)<br>9.14 (7.92 – 10.43)<br>5.28 (3.59 – 7.37) | 8.30 (7.03 – 9.83)<br>5.87 (4.68 – 7.42)<br>2.43 (1.63 – 3.39) |
| - High school | Total cause of death<br>Natural<br>External | 12.06 (10.82 – 13.49)<br>7.53 (6.48 – 8.74)<br>4.52 (3.31 – 6.14) | 4.80 (4.01 – 5.68)<br>2.84 (2.19 – 3.51)<br>1.96 (1.25 – 2.81) |
| - University | Total cause of death<br>Natural<br>External | 10.63 (8.20 – 14.14)<br>3.96 (2.49 – 5.39)<br>6.66 (3.57 – 11.09) | 1.44 (0.53 – 2.37)<br>-0.22 (-1.09 – 0.65)<br>1.66 (1.08 – 2.29) |
| Depression | | | |
| | | Inpatient | Non-inpatient |
| - Lower than high school | Total cause of death<br>Natural<br>External | 15.94 (14.31 – 18.11)<br>8.68 (7.37 – 10.04)<br>7.27 (5.22 – 10.21) | 4.14 (3.86 – 4.45)<br>3.34 (3.08 – 3.66)<br>0.79 (0.65 – 0.95) |
| - High school | Total cause of death<br>Natural<br>External | 11.11 (10.50 – 11.83)<br>6.79 (6.21 – 7.31)<br>4.32 (3.65 – 5.18) | 1.23 (1.06 – 1.39)<br>0.83 (0.68 – 0.99)<br>0.40 (0.28 – 0.51) |
| - University | Total cause of death<br>Natural<br>External | 11.84 (9.01 – 16.03)<br>3.15 (1.76 – 4.36)<br>8.69 (5.00 – 14.06) | -0.63 (-0.87 – -0.34)<br>-0.98 (-1.18 – -0.77)<br>0.35 (0.19 – 0.58) |

b) Men

| Schizophrenia | | | |
|---|---|---|---|
| | | Inpatient | Non-inpatient |
| - Lower than high school | Total cause of death<br>Natural<br>External | 19.86 (18.34 – 21.36)<br>12.04 (10.53 – 13.75)<br>7.82 (5.42 – 10.33) | 14.41 (12.67 – 16.54)<br>7.78 (6.15 – 9.79)<br>6.62 (4.66 – 9.00) |
| - High school | Total cause of death<br>Natural<br>External | 19.84 (17.42 – 22.60)<br>11.62 (9.04 – 14.28)<br>8.22 (4.79 – 12.56) | 10.44 (9.28 – 11.66)<br>5.48 (4.47 – 6.57)<br>4.96 (3.66 – 6.48) |
| - University | Total cause of death<br>Natural<br>External | 15.80 (13.18 – 19.05)<br>10.18 (7.19 – 13.92)<br>5.62 (3.49 – 8.12) | 7.59 (5.42 – 10.75)<br>3.55 (1.60 – 5.27)<br>4.04 (1.55 – 8.03) |
| Bipolar disorder | | | |
| | | Inpatient | Non-inpatient |
| - Lower than high school | Total cause of death<br>Natural<br>External | 16.20 (13.82 – 18.71)<br>8.76 (6.45 – 11.14)<br>7.44 (4.27 – 11.23) | 9.65 (8.58 – 10.79)<br>6.32 (5.22 – 7.47)<br>3.33 (2.33 – 4.50) |
| - High school | Total cause of death<br>Natural<br>External | 15.45 (12.47 – 20.12)<br>4.70 (2.36 – 6.70)<br>10.75 (6.43 – 17.70) | 5.85 (4.55 – 7.49)<br>2.27 (1.41 – 3.18)<br>3.58 (2.23 – 5.67) |
| - University | Total cause of death<br>Natural<br>External | 14.28 (11.13 – 18.13)<br>1.60 (-0.29 – 3.31)<br>12.67 (8.12 – 17.94) | 2.66 (1.67 – 3.72)<br>0.60 (-0.33 – 1.63)<br>2.06 (1.26 – 3.00) |
| Depression | | | |
| | | Inpatient | Non-inpatient |
| - Lower than high school | Total cause of death<br>Natural<br>External | 17.25 (15.93 – 18.67)<br>6.26 (5.22 – 7.31)<br>10.99 (8.96 – 13.11) | 7.31 (6.52 – 8.56)<br>3.70 (3.15 – 4.13)<br>3.60 (2.58 – 5.26) |
| - High school | Total cause of death<br>Natural<br>External | 16.32 (14.58 – 18.33)<br>6.59 (4.90 – 8.78)<br>9.73 (7.60 – 12.27) | 3.59 (3.26 – 4.00)<br>1.90 (1.58 – 2.26)<br>1.69 (1.40 – 1.99) |
| - University | Total cause of death<br>Natural<br>External | 15.05 (11.92 – 19.54)<br>2.80 (0.79 – 4.58)<br>12.26 (7.78 – 18.79) | 1.50 (0.36 – 3.34)<br>-0.36 (-0.90 – 0.16)<br>1.87 (0.60 – 4.19) |

In parentheses: 95% CI based on 1000 bootstrap iterations.

**Supplementary table 2.** Excess life-years lost from age 30 in women and men with depression in outpatient care and in primary care only, according to educational attainment.

| | | Outpatient care | Primary care only |
|---|---|---|---|
| Women with depression | | | |
| - Lower than high school | Total cause of death<br>Natural<br>External | 5.09 (4.65 – 5.57)<br>4.39 (3.98 – 4.80)<br>0.70 (0.48 – 1.01) | 4.04 (3.74 – 4.43)<br>3.21 (2.91 – 3.59)<br>0.84 (0.66 – 1.04) |
| - High school | Total cause of death<br>Natural<br>External | 2.69 (2.29 – 3.14)<br>2.00 (1.67 – 2.35)<br>0.69 (0.40 – 1.07) | 0.94 (0.76 – 1.12)<br>0.62 (0.45 – 0.78)<br>0.32 (0.21 – 0.44) |
| - University | Total cause of death<br>Natural<br>External | 0.58 (0.00 – 1.20)<br>-0.04 (-0.56 – 0.43)<br>0.62 (0.26 – 1.11) | -0.99 (-1.27 – -0.70)<br>-1.25 (-1.51 – -0.99)<br>0.26 (0.09 – 0.50) |
| Men with depression | | | |
| - Lower than high school | Total cause of death<br>Natural<br>External | 7.03 (6.27 – 7.87)<br>4.69 (4.04 – 5.31)<br>2.33 (1.58 – 3.29) | 7.43 (6.47 – 8.72)<br>3.59 (3.09 – 4.07)<br>3.84 (2.62 – 5.56) |
| - High school | Total cause of death<br>Natural<br>External | 4.48 (3.99 – 5.00)<br>2.44 (1.98 – 2.95)<br>2.04 (1.65 – 2.45) | 3.36 (2.96 – 3.85)<br>1.79 (1.45 – 2.20)<br>1.57 (1.25 – 1.95) |
| - University | Total cause of death<br>Natural<br>External | 1.69 (0.86 – 2.70)<br>0.06 (-0.49 – 0.56)<br>1.63 (0.81 – 2.84) | 1.35 (0.02 – 3.55)<br>-0.50 (-1.14 – 0.08)<br>1.85 (0.42 – 4.58) |

In parentheses: 95% CI based on 1000 bootstrap iterations.

**Supplementary table 3.** Excess life-years lost from age 30 in women and men with depression, bipolar disorder, and schizophrenia.

| | Excess life-years lost, total (95% CI) | Excess life-years lost, natural causes (95% CI) | Excess life-years lost, external causes (95% CI) |
|---|---|---|---|
| Women | | | |
| Depression | 3.11 (2.97 – 3.27) | 2.16 (2.05 – 2.28) | 0.95 (0.83 – 1.08) |
| Bipolar disorder | 8.09 (7.61 – 8.59) | 5.02 (4.61 – 5.44) | 3.07 (2.63 – 3.63) |
| Schizophrenia | 12.73 (12.04 – 13.54) | 8.55 (7.96 – 9.08) | 4.18 (3.34 – 5.20) |
| Men | | | |
| Depression | 5.89 (5.51 – 6.41) | 2.78 (2.55 – 3.00) | 3.11 (2.62 – 3.78) |
| Bipolar disorder | 9.29 (8.63 – 9.99) | 4.29 (3.82 – 4.76) | 4.99 (4.24 – 5.90) |
| Schizophrenia | 15.45 (14.59 – 16.38) | 9.08 (8.30 – 9.87) | 6.38 (5.32 – 7.54) |

In parentheses: 95% CI based on 1000 bootstrap iterations.

**Supplementary table 4.** Excess life-years lost from age 30 in women and men with depression, bipolar disorder, and schizophrenia according to level of healthcare utilization.

| | Excess life-years lost, total (95% CI) | Excess life-years lost, natural causes (95% CI) | Excess life-years lost, external causes (95% CI) |
|---|---|---|---|
| Women | | | |
| Depression, Inpatient | 13.10 (12.09 – 14.35) | 6.74 (6.06 – 7.34) | 6.36 (5.03 – 8.21) |
| Depression, Non-inpatient | 1.75 (1.62 – 1.87) | 1.27 (1.16 – 1.38) | 0.48 (0.39 – 0.57) |
| Bipolar disorder, Inpatient | 12.63 (11.61 – 13.80) | 7.20 (6.43 – 7.91) | 5.43 (4.15 – 7.05) |
| Bipolar disorder, Non-inpatient | 5.07 (4.51 – 5.71) | 3.06 (2.57 – 3.58) | 2.01 (1.61 – 2.48) |
| Schizophrenia, Inpatient | 16.55 (15.66 – 17.54) | 12.38 (11.50 – 13.33) | 4.17 (3.06 – 5.30) |
| Schizophrenia, Non-inpatient | 9.76 (8.76 – 11.01) | 5.66 (4.98 – 6.39) | 4.10 (2.93 – 5.51) |
| Men | | | |
| Depression, Inpatient | 16.28 (15.24 – 17.53) | 5.70 (4.75 – 6.71) | 10.58 (9.21 – 12.38) |
| Depression, Non-inpatient | 4.24 (3.82 – 4.84) | 1.94 (1.69 – 2.17) | 2.30 (1.81 – 3.05) |
| Bipolar disorder, Inpatient | 15.35 (13.70 – 17.56) | 5.22 (3.83 – 6.49) | 10.13 (7.84 – 13.22) |
| Bipolar disorder, Non-inpatient | 6.02 (5.35 – 6.86) | 2.95 (2.39 – 3.55) | 3.06 (2.31 – 3.95) |
| Schizophrenia, Inpatient | 19.44 (18.09 – 20.86) | 11.82 (10.43 – 13.39) | 7.63 (5.67 – 9.97) |
| Schizophrenia, Non-inpatient | 11.73 (10.74 – 12.93) | 6.21 (5.30 – 7.16) | 5.52 (4.44 – 6.93) |

In parentheses: 95% CI based on 1000 bootstrap iterations.

**Supplementary table 5.** Excess life-years lost from age 30 in women and men with depression, bipolar disorder, schizophrenia, according to educational attainment.

| | Excess life-years lost, total (95% CI) | Excess life-years lost, natural causes (95% CI) | Excess life-years lost, external causes (95% CI) |
|---|---|---|---|
| General population | | | |
| Women | | | |
| University | -2.57 (-2.67 – -2.47) | -2.44 (-2.53 – -2.34) | -0.13 (-0.17 – -0.09) |
| High school | -0.59 (-0.67 – -0.52) | -0.57 (-0.64 – -0.51) | -0.02 (-0.06 – 0.01) |
| Lower | 2.70 (2.59 – 2.81) | 2.37 (2.27 – 2.46) | 0.34 (0.27 – 0.40) |
| Men | | | |
| University | -3.19 (-3.29 – -3.09) | -2.68 (-2.77 – -2.60) | -0.51 (-0.57 – -0.45) |
| High school | -0.35 (-0.43 – -0.27) | -0.28 (-0.35 – -0.21) | -0.07 (-0.13 – -0.01) |
| Lower | 3.82 (3.67 – 3.98) | 2.75 (2.63 – 2.87) | 1.07 (0.92 – 1.24) |
| Depression | | | |
| Women | | | |
| University | 0.54 (0.25 – 0.84) | -0.31 (-0.52 – -0.10) | 0.85 (0.61 – 1.13) |
| High school | 2.49 (2.32 – 2.65) | 1.73 (1.59 – 1.88) | 0.76 (0.64 – 0.88) |
| Lower | 5.78 (5.46 – 6.12) | 4.34 (4.06 – 4.64) | 1.44 (1.23 – 1.67) |
| Men | | | |
| University | 3.14 (2.05 – 5.03) | 0.43 (-0.18 – 0.94) | 2.71 (1.42 – 5.13) |
| High school | 5.19 (4.82 – 5.60) | 2.78 (2.47 – 3.15) | 2.41 (2.12 – 2.76) |
| Lower | 8.95 (8.24 – 10.03) | 4.41 (3.91 – 4.82) | 4.53 (3.56 – 6.08) |
| Bipolar disorder | | | |
| Women | | | |
| University | 4.77 (3.88 – 5.78) | 1.73 (1.09 – 2.35) | 3.04 (2.17 – 4.27) |
| High school | 7.82 (7.17 – 8.53) | 4.93 (4.42 – 5.47) | 2.89 (2.22 – 3.64) |
| Lower | 10.94 (9.96 – 12.05) | 7.63 (6.70 – 8.84) | 3.31 (2.57 – 4.15) |
| Men | | | |
| University | 6.54 (5.57 – 7.69) | 1.82 (1.06 – 2.60) | 4.72 (3.47 – 6.04) |
| High school | 9.29 (8.16 – 10.73) | 3.78 (2.95 – 4.55) | 5.51 (4.08 – 7.46) |
| Lower | 12.00 (10.96 – 13.11) | 7.42 (6.40 – 8.51) | 4.57 (3.46 – 5.98) |
| Schizophrenia | | | |
| Women | | | |
| University | 8.96 (6.74 – 12.29) | 3.73 (2.28 – 5.27) | 5.23 (2.66 – 9.27) |
| High school | 11.78 (10.79 – 12.85) | 7.50 (6.62 – 8.40) | 4.28 (3.10 – 5.56) |
| Lower | 14.66 (13.74 – 15.64) | 10.91 (10.01 – 11.81) | 3.76 (2.78 – 4.85) |
| Men | | | |
| University | 11.25 (9.45 – 13.14) | 6.44 (4.81 – 8.27) | 4.82 (2.99 – 7.23) |
| High school | 14.81 (13.63 – 16.11) | 8.60 (7.50 – 9.75) | 6.20 (4.65 – 8.08) |
| Lower | 17.29 (16.10 – 18.76) | 10.07 (8.79 – 11.50) | 7.21 (5.65 – 9.07) |

In parentheses: 95% CI based on 1000 bootstrap iterations.
